# Diagnosis and Prediction Model for COVID-19 Patient’s Response to Treatment based on Convolutional Neural Networks and Whale Optimization Algorithm Using CT Images

**DOI:** 10.1101/2020.04.16.20063990

**Authors:** Sally Elghamrawy, Aboul Ella Hassanien

## Abstract

The outbreak of coronavirus diseases (COVID-19) has rabidly spread all over the world. The World Health Organization (WHO) has announced that coronavirus COVID-19 is an international pandemic. The Real-Time Reverse transcription-polymerase Chain Reaction (RT-PCR) has a low positive and sensitivity rate in the early stage of COVID-19. As a result, the Computed Tomography (CT) imaging is used for diagnosing. COVID-19 has different key signs on a CT scan differ from other viral pneumonia. These signs include ground-glass opacities, consolidations, and crazy paving. In this paper, an Artificial Intelli-gence-inspired Model for COVID-19 Diagnosis and Prediction for Patient Response to Treatment (AIMDP) is proposed. AIMDP model has two main functions reflected in two proposed modules, namely, the Diagnosis Module (DM) and Prediction Module (PM). The Diagnosis Module (DM) is proposed for early and accurately detecting the patients with COVID-19 and distinguish it from other viral pneumonias using COVID-19 signs obtained from CT scans. The DM model, uses Convolutional Neural Networks (CNNs) as a Deep learning technique for segmentation, can process hundreds of CT images in seconds to speed up diagnosis of COVID-19 and contribute in its containment. In addition, some countries haven’t the ability to provide all patients with the treatment and intensive care services, so it will be mandatory to give treatment to only responding patients. In this context, the Prediction Module (PM) is proposed for predicting the ability of the patient to respond to treatment based on different factors e.g. age, infection stage, respiratory failure, multi-organ failure and the treatment regimens. PM implement the Whale Optimization Algorithm for selecting the most relevant patient’s features. The experimental results show promising performance for the proposed diagnosing and prediction modules, using a dataset with hundreds of real data and CT images.

## 1 Introduction

. On December 2019, a severe acute respiratory syndrome coronavirus 2 (SARS-CoV-2), named as COVID-19 by the World Health Organization (WHO) on February 2020 [1], occurred in the city of Wuhan in China [2,3]. An outbreak of coronavirus diseases (COVID-19) has rabidly spread all over the world. The World Health Organization (WHO) has announced that coronavirus COVID-19 is an international pandemic.

To the date (Mar 28th 2020), there have been 597,458 confirmed cases and 27,370 deaths all around the world [4]. COVID-19 is extremely contagious and easily spreadable between people via respiratory droplets. For mild cases, it may cause some common symptoms as shortness of breath, muscle pain, fever and sputum production. While for severe cases, it may lead to some progress to pneumonia and multi-organ failure [5-7]. Unfortunately, the death rates/ diagnosed cases number recorded on March 2020 is 4.5%. Many countries have been affected, and there are enormous cases of community spread, meaning that individuals are getting infected without contacting sick persons or travelling to infected areas.

With the daily rapid growth in the number of newly confirmed and suspected cases, the diagnosis has become a critical issue that effect the containment of disease, especially in the epidemic countries, that suffer from lack of resources and low detection rates [8]. In this context, healthcare workers need sensitive and specific diagnosing tools to investigate cases of potential COVID19. Many researchers [9-12] have been developing many tools using Artificial Intelligent techniques and machine learning in building response prediction and diseases diagnosis models for early detection of COVID 19 to contribute in its containment before spreading. These models assist clinicians in making the appropriate recommendations for treatments.

The gold standard for diagnosis of COVID-19 is the Reverse Transcription Polymerase Chain Reaction RT-PCR [13]. Unfortunately, current data suggest that RT-PCR is initially only 30–70% sensitive for acute infection. In other words, 3 to 7 out of 10 patients that actually have COVID-19 will have a negative PCR result. However, a newest study of over 1000 patients [11-14] showed that chest Computed Tomography CT has an initial sensitivity of over 95%, meaning that fewer than 1 out of 20 cases would get missed. That has massive implications because far fewer people with COVID-19 would be sent back home where they could get sicker, and spread the disease to others. However, number of radiologists stated that CT should be used sparingly in COVID-19 and only when impacting management. For that reason, a supervised computer-aided CT diagnosis models are immediately needed for accurately detecting COVID-19 cases.

In this Paper, an Artificial Intelligence-inspired Model for COVID-19 Diagnosis and Prediction for Patient Response to Treatment (AIMDP) is proposed, which can automatically select and detect the COVID-19 signs (features) from CT images. AIMDP has two main modules: The Diagnosis Module (DM) and the Prediction Module (PM). The Diagnosis Module (DM) in AIMDP uses CT images and further lab evaluation, when needed, to early and accurately diagnosing cases with COVID-19. The DM model, uses Convolutional Neural Networks (CNNs) as a Deep learning technique for segmentation, can process hundreds of CT images in seconds to speed up diagnosis of COVID-19 and contribute in its containment. To accurately detect the signs of COVID-19 in CT images, a Feature Selection phased bases on Whale Optimization Algorithm (FSWOA) is proposed for selecting the most relevant patient’s features. The most common signs for COVID 19, detected on CT scans are the Ground Glass Opacities 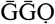, that represent tiny air sacs (alveoli) filling with fluid and turning a shade of grey in CT scan. In severe infections and more advanced infections, more fluid will be occurring in lobes of the lungs, so the ground glass opacities will progress to Solid White Consolidation 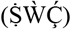 sign. However, the Crazy Paving Pattern 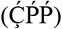 swelling due the swelling in interstitial space along the walls of the lungs that makes the wall looks thicker like the white lines against the hazy ground glass background grey. The 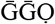 is usually the first sign of COVID19, and followed later by one or both of the other signs. Moreover, some countries haven’t the ability to provide all patients with the treatment and intensive care service, it will be mandatory to give treatment to only responding patients. In this context, the Prediction Module (PM) is proposed for predicting the ability of the patient to respond to treatment based on different factors e.g. age, infection stage, respiratory failure, multi-organ failure and the treatment regimens. PM implement the Whale Optimization algorithm for selecting the most relevant patient’s features. The experimental results show promising performance for the proposed diagnosing and prediction modules, using a dataset with hundreds of real data and CT images.The rest of the paper is organized as follows: Section 2 shows the recent COVID-19 diagnosing models proposed, and a background of WOA is introduced. Section 3 demonstrates the proposed Artificial Intelligence-inspired Model for COVID-19, the Diagnosis (DM) and the Prediction Module (PM) are also discussed in details. The performance of AIMDP is evaluated in section 4 showing the effect of implementing FSWOA and using CNNs as a deep learning technique. The results are compared against the most recent diagnosing models’.

### 1.1 Main Contributions of the Proposed AIMDP Model

Thus, the development of AI techniques to be used in diagnosing models will be a great help to healthcare workers. In this context, AIMDP model is proposed to utilize different AI techniques to enhance the diagnosis and prediction function of the model as follows:

First, in the segmentation phase, before using CNNs as a deep learning technique for segmenting the CT images, a pre-processing phase is implemented to only consider the lung regions and remove the noise in the non-lung regions to avoid time consuming in segmenting the whole image. It also normalizes the image and assign a label for each patch/image to represent each feature detected.

Second a Feature selection phase is used to select the minimum set of relevant features of diagnosing COVID-19 using WOA to optimize the results obtained.

Third, the classification phase uses different classifiers and the model choose the most appropriate classifier based on the classification error obtained for each case.

Even though chest CT is very sensitive to COVID19, there are some COVID 19 key findings like (Ground Glass Opacities, Solid White Consolidation and Crazy Paving Pattern) that also can be seen in other causes of Viral pneumonias like influenza and adenovirus. That means, the chest CT scan images can be sensitive but not specific for COVID19. For this reason, in the diagnosing and classification phase of AIMDP, further evaluation from lab tests (RT-PCR and CBC) are used to exclude other causes and accurately diagnosis COVID-19, and doesn’t only depend on CT images for diagnosing Finally, AIMDP has a prediction phase that gives a probability of patient ability to respond to the COVID-19 treatment based on different inputs given for the patient like his age, infection stage, respiratory failure, multi-organ failure and the treatment regimens. This predicted value is given to the doctors that make a decision to provide the patient with treatment and intensive care services. This will help the doctors to save the resources of endemic countries, that have no enough resources, and only give the treatment to the responding patients.

## 2 Related Works

Due to the rapid expansion of AI technology that has been commonly applied in the medical field, different studies were conducted on the diagnosis and classification of different diseases like viral pneumonias and organs’ tumours. Nowadays, due to the COVID-19 spreading disaster, many researches have focused on diagnosing and detecting COVID-19 as follows: Authors in [9] proposed a 3D deep convolutional neural Network to Detect COVID-19 from CT volume, namely DeCoVNet. But, the algorithm worked in a black-box manner when diagnosing COVID-19, since the algorithm was based on deep learning and its explain ability was still at an early stage. COVNET [10] developed a framework to detect COVID-19 using chest CT and evaluate its performances. The authors proposed a three-dimensional deep learning framework to detect COVID-19 using chest CT. Community acquired pneumonia (CAP) and other nonpneumonia exams were included to test the robustness of the model.

In addition, Yang et al. [11] investigated the diagnostic value and consistency of chest CT as compared with comparison to RT-PCR assay in COVID-19. Their analysis suggests that chest CT should be considered for the COVID-19 screening, comprehensive evaluation, and following-up, especially in epidemic areas with high pre-test probability for disease. Jiang et al [12] proposed established an early screening model to distinguish COVID-19 pneumonia from Influenza-A viral pneumonia and healthy cases with pulmonary CT images using deep learning techniques. The authors used multiple CNN models to classify CT image datasets and calculate the infection probability of COVID-19. The findings might greatly assist in the early screening of patients with COVID-19 by deep learning technologies. The authors proposed a location-attention mechanism and uses it in the classical ResNet for feature extraction. The authors in [13] constructed a system based on deep learning for identification of viral pneumonia on CT.

Although, these proposed models can result acceptable outcomes, their executions include many procedures which may need longer time. Moreover, using the deep learning methods in diagnosing is time consuming and difficult for radiologists, especially when there are thousands of images to be processed huge amount. In addition, the models that depend on deep learning techniques only leads to black box problem when diagnosing COVID-19. In this context, AIMDP model avoid these drawbacks, by utilizing different AI techniques to enhance the diagnosis and prediction function of the model.

### 2.1 Whale Optimization Algorithm WOA

WOA is a is one of the latest nature-inspired metaheuristic algorithm introduced by Mirjalili [15], WOA emulates the social behaviour of humpback whale when hunting the prey. A detailed descriptions of the whales’ behaviours can be found in [16]. The Humpback whales hunt group of krill (preys), which closes to the surface by spinning around them within a shrinking circle and creating bubbles along a ‘9’-shaped path, this technique is called spiral bubble-net feeding method, as shown in figure 1. The spiral bubble-net attacking method were represented in the exploitation phase, while the random search process for the preys were represented in the exploration phase, the following subsections discuss the mathematical model of each phase.

**Figure 1.**
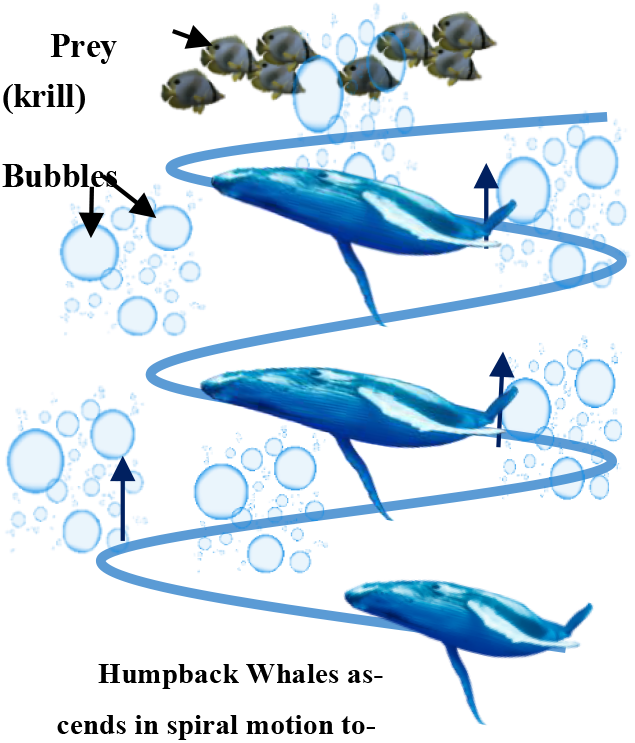
The spiral bubble-net attacking method of humpback whales.

#### The Spiral Bubble-Net Attacking Method

For encircling the prey, number of whales are updating their positions towards the best position of the prey. These actions are described as in (1) and (2)

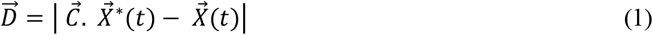

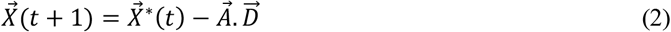

Where (t) that specifies the current iteration, 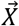 is the position vector, 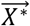 represents the best solution obtained so far, 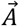 *and* 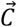 are coefficient vectors that are calculated as in (3) and (4), respectively:

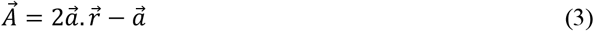

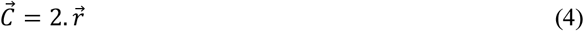

Where 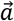 is linearly decreased from 2 to 0 over the course of iterations in exploration and exploitation phases, and 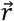 is a random vector generated with uniform distribution in the interval of [0,1].

In the exploitation phase, WOA algorithm updates the position of whale during optimization by choosing between either the shrinking encircling mechanism or the spiral updating position model.

#### The Global Search for the Pray

In the exploration phase, the humpback whale searches for the prey randomly. A random search agent is selected to guide the search and the other search agents update their positions based on this selected agent, instead of depending on the optimum search agent found so far. So, if the 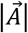 has the random values greater than 1, this will enforce search agent to move far away from the best-known search agent as in (7) and (8):

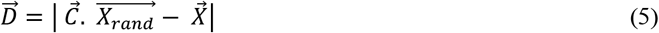

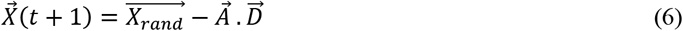

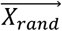 is a random position for the randomly chosen whale from the current population of whales. In WOA, the positions of the search agents are updated at each iteration according to the value of 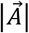, if 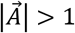 the position will be updated randomly according to randomly chosen search agent and if 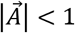, the position will be updated according to the best solution. So, the parameter (a) is used to switch smoothly between exploration and exploitation phases. Also, the parameter *p* controls the switching between the two kinds of whale’s movement “spiral or circular motion.

## 3 The Proposed Artificial Intelligence-inspired Model for COVID-19 Diagnosis and Prediction for Patient Response to Treatment (AIMDP)

In the proposed model, different AI techniques are used based on their functionality on six main phases, as shown in figure 2, namely, the pre-processing, segmentation, feature selection, classification, Prediction and diagnosis recommendation phase. The AIMDP model has two main modules: The Diagnosing Module (DM) which is concerned with detecting the patients with Coronavirus (COVID 19) Disease and distinguish it from other viral pneumonias, by detecting the most relevant features using feature selection and classification phases. After the patient has been detected as a COVID 19 case, the doctors must decide which patient will be provided with treatment and intensive care service. It is worth mentioned that not all cases have the same response to treatment. When there is a lack of resources, it will be mandatory to give treatment to only responding patients. For this reason, the Prediction Module(PM) is used to predict different patients’ responses to COVID 19 treatment. And based on this prediction, the doctor will decide the best treatment regime for patients. Considering the lack of resources and the gradually growth of COVID19 cases, an early assessment of the patient response to treatment is essential, especially for non-responders. In addition, if such non-responders could be predicted early, an alternative regime will be studied and considered for them.

**Figure 2.**
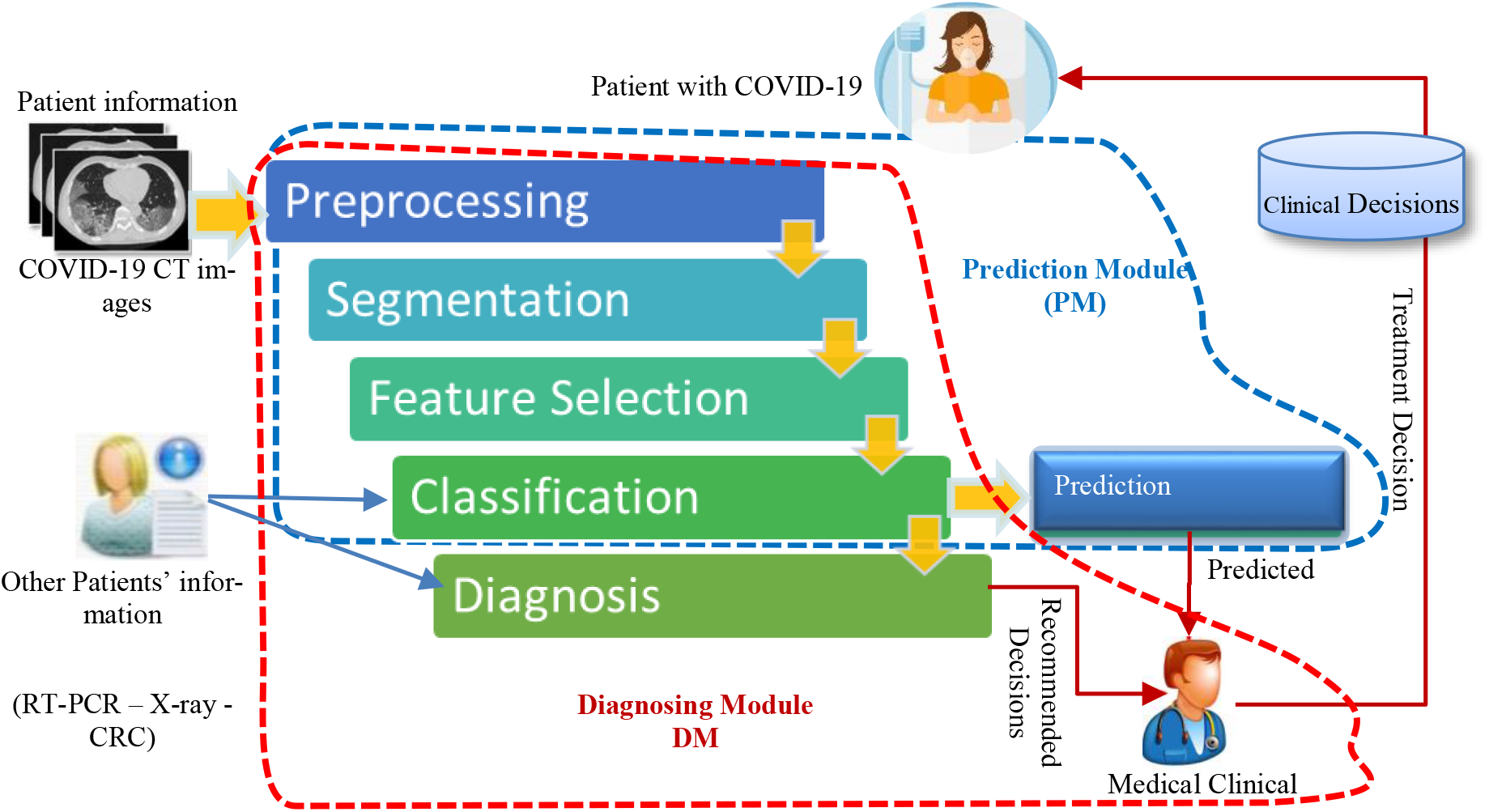
The flow diagram of the Proposed Artificial Intelligence-inspired Model of Diagnosing and Prediction COVID-19 cases

The detailed description of the proposed AIMDP model is shown in figure 3. First the model starts by taking the patient information as the input; this information contains the raw CT images data about suspected patients. The pre-processing phase handle any noise or missing data that might occurs in the original dataset. The CT images in classes are then sorted based on their features and attributes to extract effective pulmonary region.

**Figure 3.**
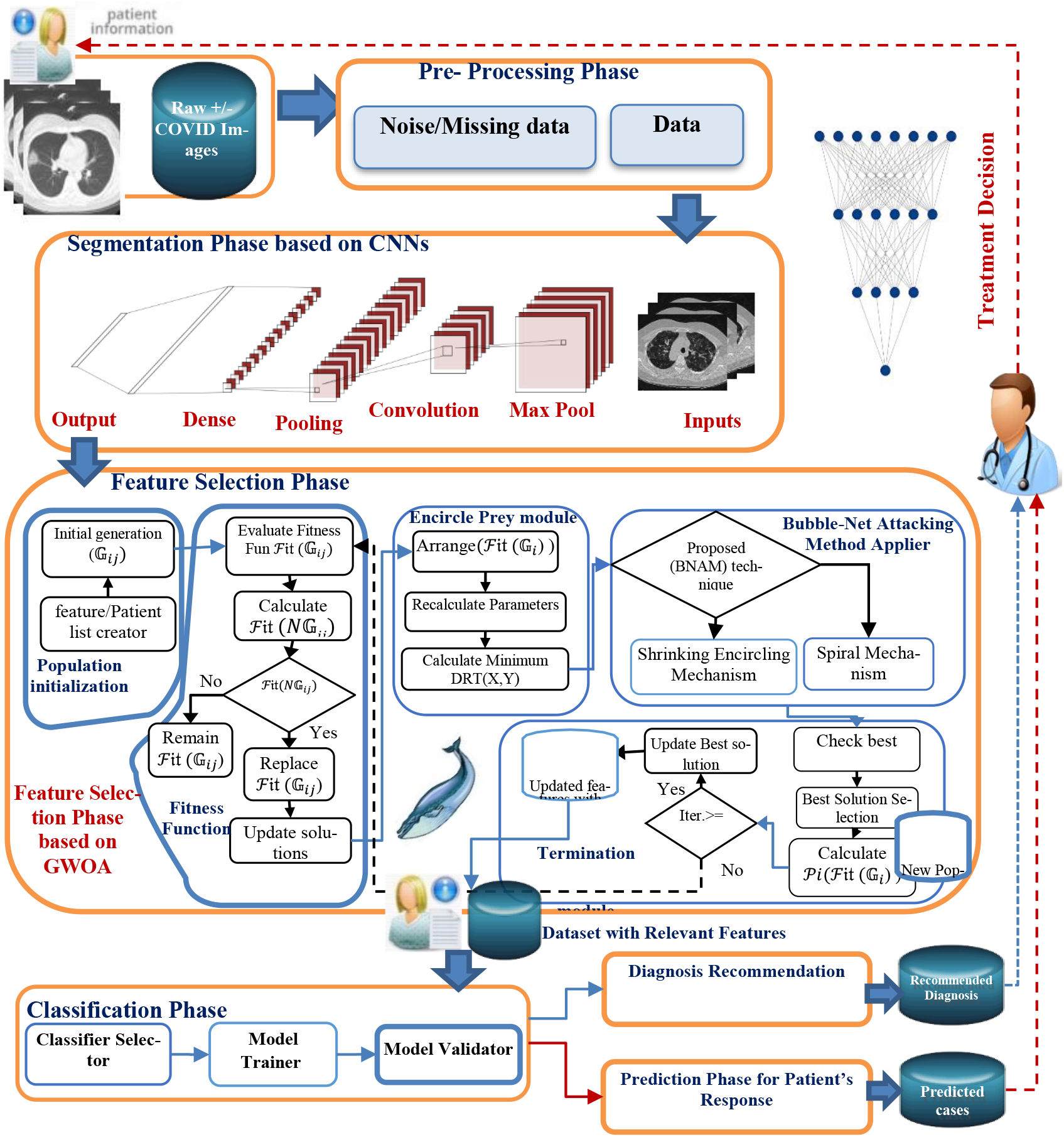
The Proposed Artificial Intelligence-inspired Model for COVID-19 Diagnosis and Prediction for Patient Response to Treatment (AIMDP)

### 3.1 Segmentation Phase

In the segmentation phase, the Convolutional Neural Networks (CNNs) is used as a deep learning technique. CNNs were initially proposed by Fukushima [17]. Different filters are applied in CNNs to capture the relevant features from image using predefined parameters and learned weights at every level. In this phase, the following sequence of layers are presented: (1) The Maximum Pooling (Max Pool) layer that is used to reduce the features in CT image by summarizing the most stimulated occurrences of a feature. (2) The convolution Layer: used to convolve a kernel(filter) of weights to extract the features. (3) Pooling Layer that use statistical data about the surrounded features to reduce the resolution. (4) The dense Layer that search for specific patterns in pixel values and classify the features with same patters in specific classes.

The segmentation phase processes hundreds of CT images in seconds to speed up diagnosis of COVID-19 and contribute in its containment. A sample of the images output from the segmentation phase is shown in figure 5.

**Figure 5.**
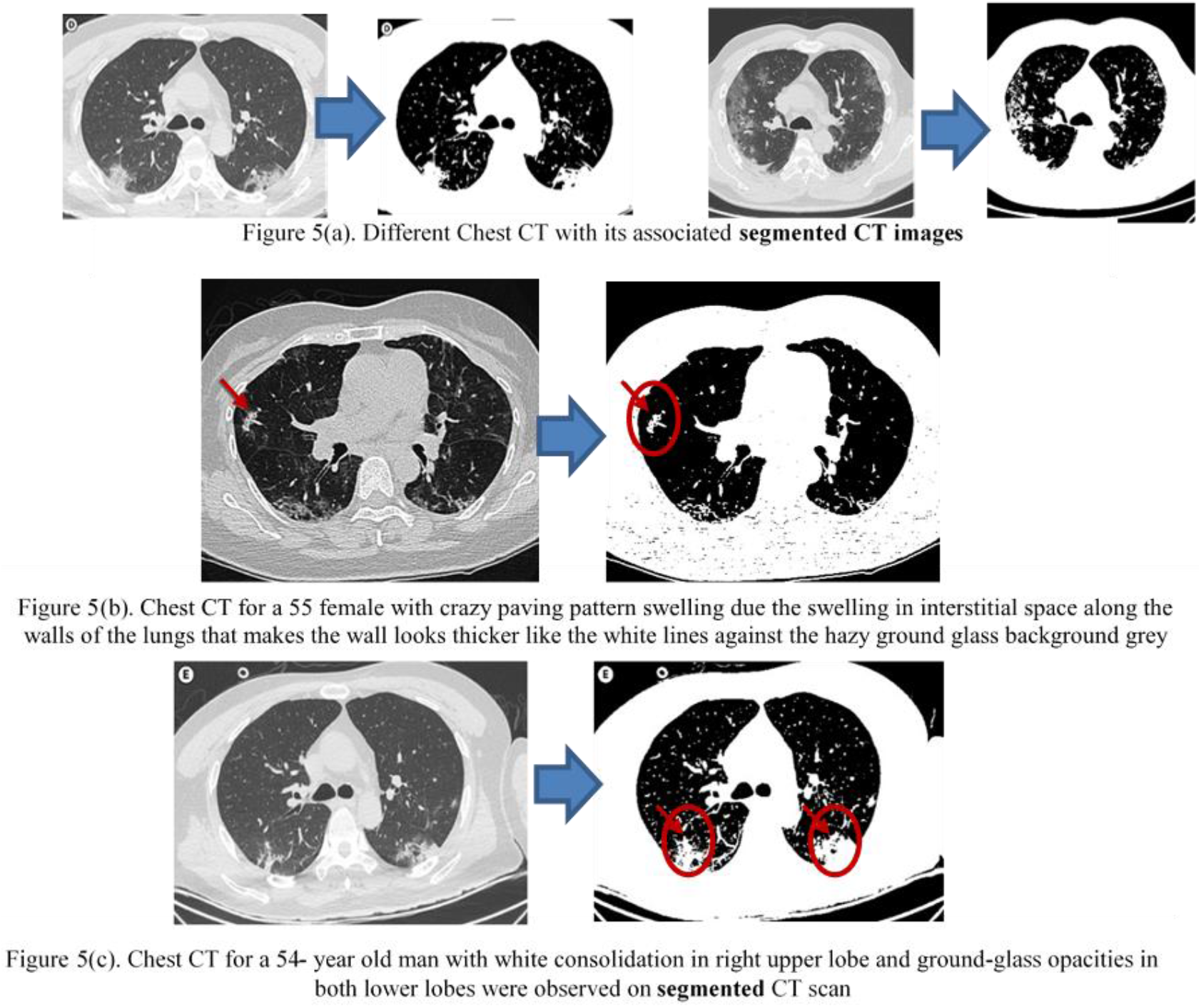
A segmented CT images output from the segmentation Phase of AIMDP.

This phase splits all CT images into patches, and then these patches are input to the trained CNN. Concurrently, a label will be assigned for each patch to represent each feature detected, as shown in figure 6. These labels are then collected to represent the CT key findings features like Ground Glass Opacities, Solid White Consolidation, Crazy Paving Pattern, Pleural Effusions, Large Lymph Nodes and Lung Cavities. The final results of segmentation of lung region are created by collecting these features.

**Figure 6.**
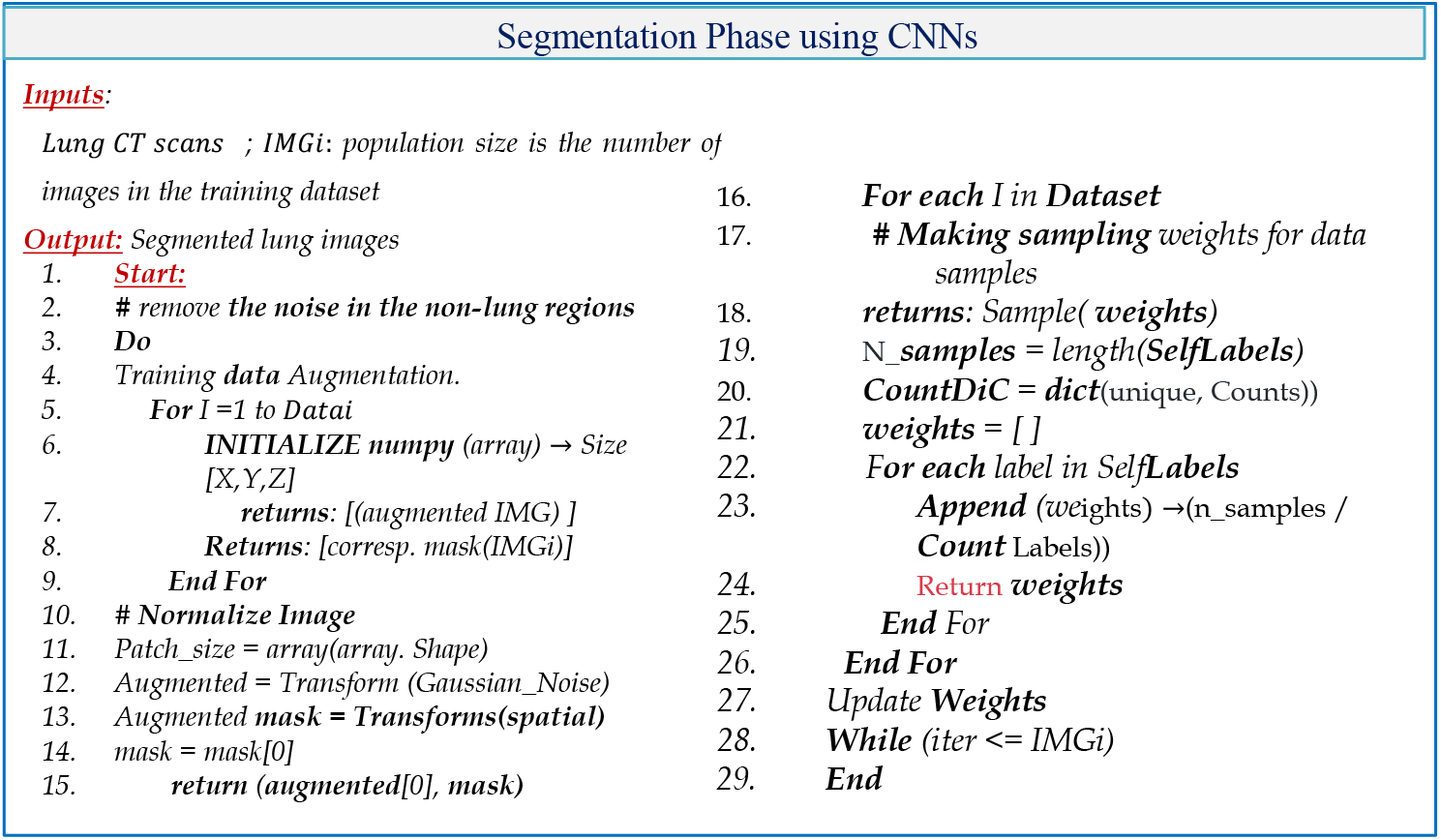
The Pseudocode for segmentation Phase: The CT images are split into patches.

### 3.2 Feature Selection (FS) Phase based on Whale Optimization Algorithm (FSWOA)

Whale Optimization Algorithm WOA is one of the latest nature-inspired metaheuristic algorithm introduced by Mirjalili [15], WOA emulates the social behaviour of humpback whale when hunting the prey. A detailed descriptions of the whales’ behaviours can be found in [16]. The feature selection phase in AIMDP use the WOA algorithm to select the most optimal relevant features, from the detailed patient information, to distinguish COVID 19 disease from different vial phonemes. The FS phase consists of five modules: Population initialization, Fitness Function, Encircle Prey, Bubble-Net Attacking Method Applier and Termination Module. In the Population initialization module, a number of search agents (whales) is randomly initialized, each whale represents a subset of features, to target the most relevant features (Preys). The appropriate representation of the feature/ patient list will help searching for the optimum solution and optimize the accuracy of diagnosing COVID19 disease. In the fitness function module, these agents(features) are converted into possible solutions for a given fitness function and initial generation of WOA parameters are set. Then for each solution, a fitness function will be evaluated for this solution. WOA uses fitness function that selects the minimal set of features for a patient achieving the best accuracy in diagnosing the virus. The most common CT finding in COVID-19 is the Ground Glass Opacities 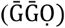 They represent tiny air sacs (alveoli) filling with fluid and turning a shade of grey in CT scan. In severe infections and more advanced infections, more fluid will be occurring in lobes of the lungs, so the ground glass opacities will progress to Solid White Consolidation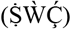.

However, the Crazy Paving Pattern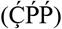 swelling due the swelling in interstitial space along the walls of the lungs that makes the wall looks thicker like the white lines against the hazy ground glass background grey. The 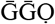 is usually the first sign of COVID19. And followed later by one or both of the other signs. It is worth mention that some CT finding usually not seen in COVID19, or seen less often, like Pleural Effusions 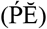, Large Lymph Nodes (LLŇ) and Lung Cavities(ĹČ) that can be devolved in other pneumonias. From these findings, a fitness function can be calculated as follows:

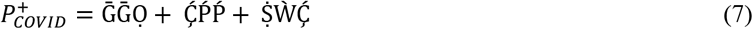

Where 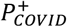 is the function used to indicate the probability of positivity of COVID19,

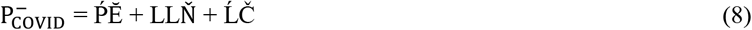

Where 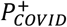 is the function used to indicate the probability of negativity of COVID19,

The fitness function for each feature(whale) position is evaluated, as follows:

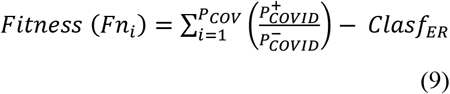

Where *P*_*COV*_ is the total number of features. *Clasf*_*ER*_ represents the classification error rate of a given classier.

After calculating the fitness function for each feature, the fitness values are compared with the previous one. If the new values are greater than old ones, then the search agents replace the old values with the new values of solutions, otherwise the old solutions are retained, as shown in figure 3.

The encircle prey module arrange the fitness values obtained then it searches for the optimum solution in the neighbour of the best-known solution using the Distance Ratio Test (DRT) technique to detect the exact location of the solution(prey) in terms of (x, y) dimensions, during the searching process of preys [18]. In the Bubble-Net Attacking Method Applier module, a Bubble-Net Attacking Method Applier (BNAM) technique is used, first proposed by [18], to switch between the attacking methods of the whale depends the location of the prey. Based on the solution’s location detected and compared with other locations, BNAM select the suitable mechanism for this location, it switches between the shrinking encircling and spiral mechanism. In the termination module, the maximum number of iterations (*Maxit*) is predefined from the beginning, and when it reached the search process will terminated. Then, the best solution is chosen which represent the highest fitness solution obtained.

### 3.3 The Classification Phase

These selected features are passed to the classification phase, that use some additional data from RT-PCR and Complete Blood Count (CBC), when needed, to accurately classify the patients based on their viral pneumonias signs and features. AIMDP model uses the classifier selector module to choose the classifier with most accurate value for the tested case. The tested classifiers are: SVM, Naive Bayes (NB), and Discriminant Analysis (DA) to test the performance with different perspectives.

In the proposed model, the FSWOA phase provides the classifier with number of whales, each whale signifies a group of features “1” means that the feature is selected and “0” means not selected. Thus, WOA search to find the most relevant set of features that achieves the highest accuracy with either SVM, NB or DA classifier and the fitness function is utilized. The outcome of the classification is estimated based on the values of optimum features obtained from CT scan. And if the case is suspected (not confirmed) as a COVID-19 case, then further lab evaluations must be considered for accurate classification. The classification phase main goal is to differentiate COVID-19 patient from other infections. After classifying the data, the model is trained and validated in this layer. A confusion matrix is produced as a graphic form of the performance. each row refers to the instances in its real class while each column refers to the instances in a predicted class. Based on this matrix, the sensitivity, specificity, accuracy and f-measure are calculated to evaluate the classifier.

### 3.4 Diagnosis Recommendation Phase

In this phase, the performance of each classifier’s prediction is evaluated based on further evaluation from lab tests, like RT-PCR and Complete Blood Count CBC, to exclude other causes and to accurately diagnosis COVID-19.

Diagnosis phase uss CT chest scans to diagnosis COVID-19 based on the relevant signs extracted. As we mentioned before that the ground glass opacities is usuall the first sign of COVID19 and it can apear isolated or combined with consolidations and cravy paving pattern. These finding usually appear in multiple lobes in both lungs and outer periphery of the lungs. In mild and recovery cases of COVID19, the sings can be isolated to be in just one lobe. The motivation of finding these three signs were be more likely to be recognized as COVID-19. Otherwise, these finding can also be seen in other causes of Viral pneumonias like influenza and adenovirus. That means, in this case the diagnosing cant depend on chest CT scan images only, as it can’t be reliable. The diagnosing phase uses further evaluation from lab tests to exclude other causes and accurately diagnosis COVID-19. The recommendations are then sent to the clinical, that must deal with the challenge of highly increasing numbers of patient and analysing high-dimensional patients’ data, to give a final decision for a specific patient.

### 3.5 Prediction Phase for Patient’s Response to COVID-19 Treatment

In some endemic countries, there are no resources or ability to provide all patients with the treatment and intensive care service. As a result, there must be a decision to be made whether to provide the treatment to the patient with highly chance to be recovered, or to provide the treatment to a non-responder patient. In this context, the prediction module, based on Whale Optimization Algorithm, is proposed for predicting the ability of the patient to respond to treatment based on different factors e.g. age, infection stage, respiratory failure, multi-organ failure and the treatment regimens. Figure 7 shows the flow diagram of diagnosis and predicting patient response. This will help the healthcare workers to search for new treatment for non-Responders, to early isolate the responders and to early provide treatment for responders.

**Figure 7.**
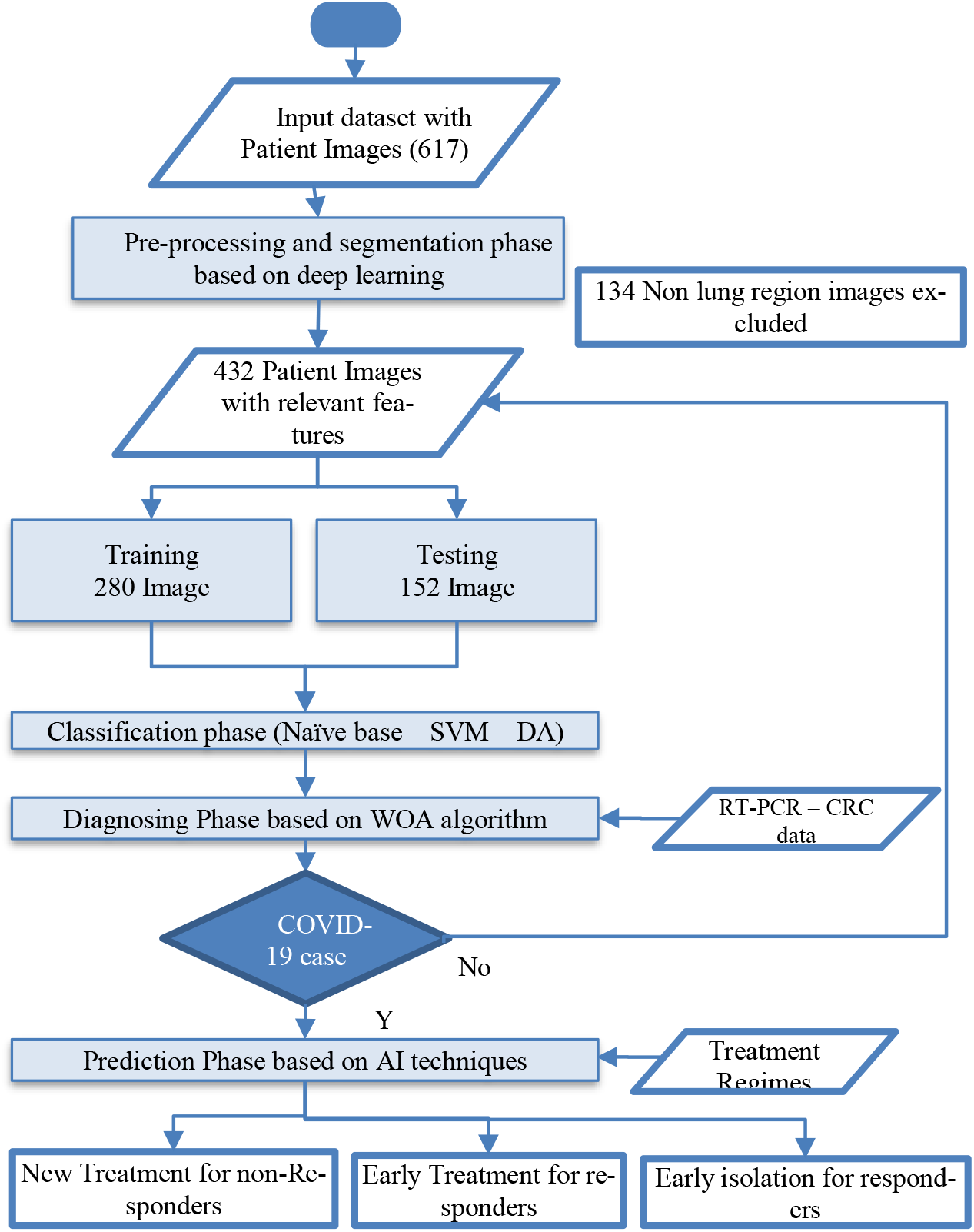
The Flow diagram for the Diagnosing and Predicting COVID-19 cases.

Different studies [19-20] had been made to study the characteristics of COVID-19 recovered cases. The studies showed that the older the patient, the more risk they face from the virus. While most of the verified cases involved people between the ages of 30 and 69, the fatality rate increased as the patients’ ages went up. For patients with 80 years old and older, 14.8% of those died compared to 0.2% for patient with 49 years or younger. In addition, the study showed that patients who suffered from chronic diseases before contracting the virus were more likely to die. These studies highlighted the importance of predicting patient response to COVID 19 to provide the treatment and intensive care services only to responding patients The disease severity is proportional to lung findings. As the patient improves, there is gradually resolution of ground glass and consolidations.

Using the Whale Optimization Algorithm (WOA) in the prediction phase of AIMDP optimizes the results obtained due to selecting the minimum set of non-responders’ patient features that are most relevant for prediction. The first data used is the data before the initiation of the treatment. The later one was done after the first cycle of the COVID-19 treatment. A set of selected features have been selected by WOA and calculated for each pateint. These selected features are: age, history in chronic diseases, respiratory failure, multi-organ failure and the treatment regimens. Finally, a PV value is calculated indicating the Prediction Value for responding patient.

## 4 Experimental Evaluation

A number of simulated experiments were performed to validate the effectiveness of the proposed model and its associated modules. The methods are implemented in MATLAB R2019a and are executed on a windows 10 PC with Intel(R) Core (TM) i7 CPU, with 16 GB RAM and 2.81 GHz clock speed. Table 1 shows the evaluation parameters used in implementing the AIMDP model.

**Table 1:** The evaluation parameters for the AIMDP model.

| Parameter | Value | Parameter | Value |
| --- | --- | --- | --- |
| Number of generations(cycle) | 300 | Number of images | 483 |
| Population size $PopS$ | 100 | Time required to train/test CNNs with all images | 244.6 / 98.94 Sec |
| Maximum number of iterations $Max_{it}$ | 10 | The average prediction time for model | 20.4 Sec/Patient |
| Threshold for CNNs | 0.4 | The K-fold Cross-validation | 10 |

A total of 617 CT scans chest were collected from different resources [21-23]. 134 Non lung region images were excluded from testing. 432 patient diagnosed with the COVID-19, 151 patients infected with other viral pneumonias.

AIMDP model uses the CNNs as a deep learning technique, the TensorFlow [19] framework is used, one of the most popular deep learning open source libraries. Figure 8 shows a snapshot of the training process on TensorFlow.

**Figure 8.**
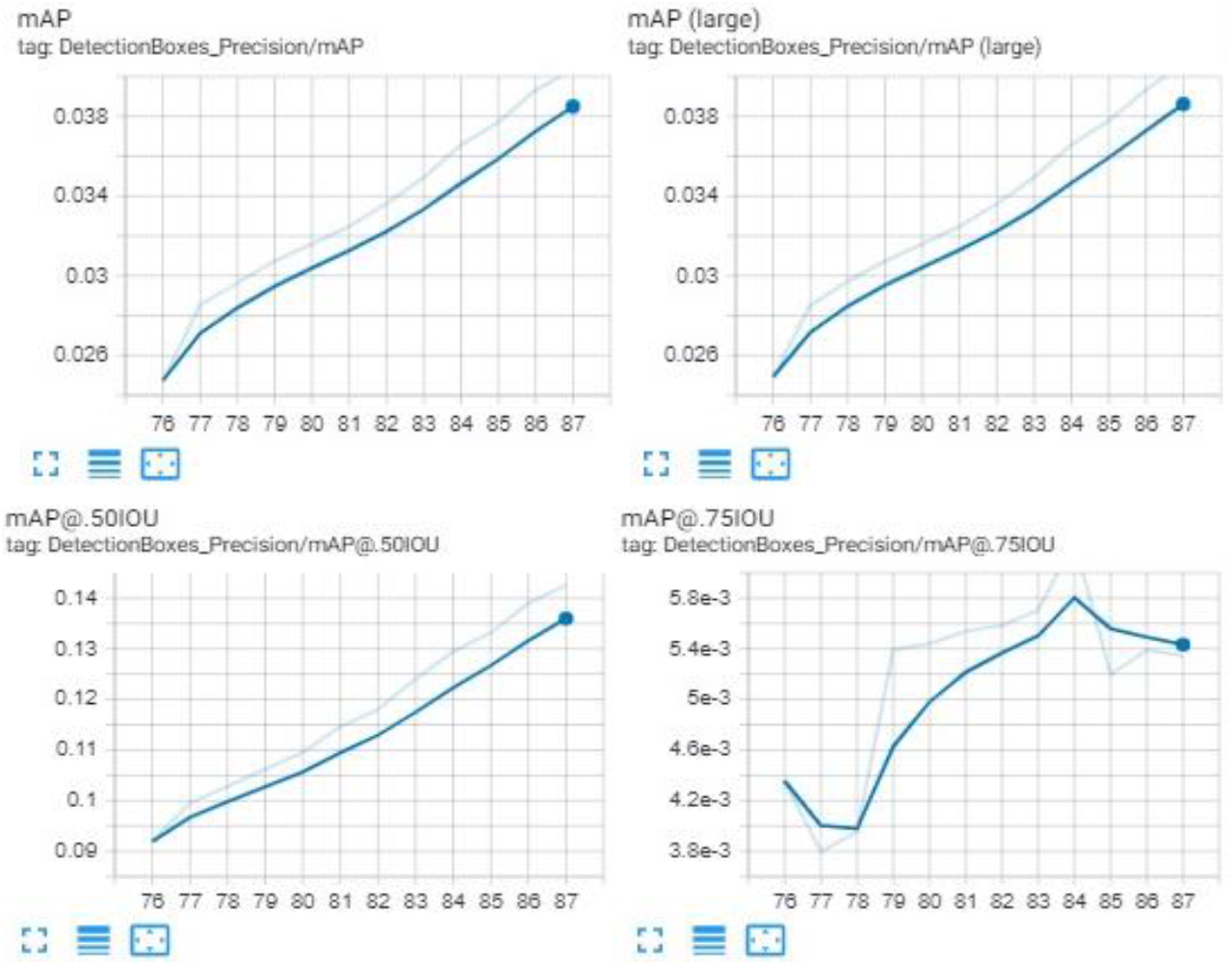
A Snapshot of the training Process using TensorFlow.

### 4.1 Performance Evaluation Measures

To evaluate the effectiveness of the proposed models, the F-measure(Score), accuracy, precision, recall(Sensitivity) and specify measures are considered. Table 2 shows the confusion matrix that used to calculate these performance, where TP, TN, FP and FN are true positive, true negative, false positive, and false negative respectively.

**Table 2.** Confusion matrix.

|  | Predicted as Positive | Predicted as Negative |
| --- | --- | --- |
| Actually Positive | True Positives (TP) | False Negatives (FN) |
| Actually Negative | False Positive (FP) | True Negatives (TN) |

**Table 2:** The overall accuracy, F-score, G-mean, and the AUC for AIMDP using different classifiers.

|  | SVM |  |  | NB |  |  | DA |  |  |
| --- | --- | --- | --- | --- | --- | --- | --- | --- | --- |
|  | Non COVID | COVID | Overall | Non COVID | COVID | Overall | Non COVID | COVID | Overall |
| **Accuracy** | 97.14 | 97.14 | 97.14 | 95.99 | 95.99 | 95.99 | 94.71 | 94.71 | 94.71 |
| **Precision** | 98.03 | 95.47 | 96.75 | 98.64 | 91.44 | 95.04 | 95.46 | 93.22 | 94.34 |
| **Sensitivity** | 97.6 | 96.27 | 96.93 | 95.2 | 97.51 | 96.35 | 96.51 | 91.29 | 93.9 |
| **F-Score** | 97.81 | 95.87 | 96.84 | 96.89 | 94.38 | 95.63 | 95.98 | 92.24 | 94.11 |
| **G-mean** | 96.93 | 96.93 | 96.93 | 96.35 | 96.35 | 96.35 | 93.86 | 93.86 | 93.86 |
| **(AUC)** | **0.99573** |  |  | 0.98572 |  |  | 0.98599 |  |  |

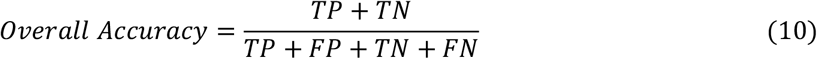

The important measures of the performance are: True Positive Rate (TPR), True Negative rate (TNR) and, Positive Predictive Value (PPV), defined as follows,

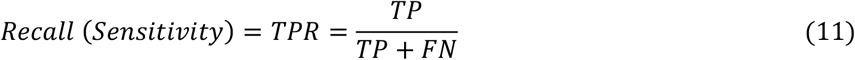

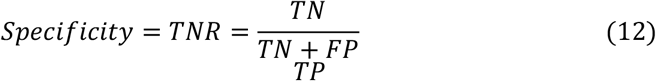

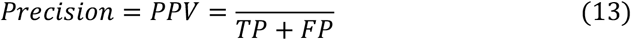

It is more difficult to obtain both high precision and high recall simultaneously in imbalanced learning. F-measure (also known as F-score) is a typical metric for the imbalanced data classification [34]. It represents a harmonic mean between recall and precision

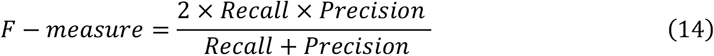

The harmonic mean of two numbers tends to be closer to the smaller of the two. Hence, a high F-measure value ensures that both recall and precision are reasonably high. The receiver operating characteristic curve, or ROC curve, is an assessment technique that makes use of the TPR and FPR. On an ROC curve, the x-axis is FP percentage, which can be calculated by the formula,

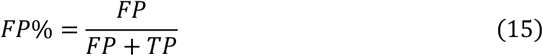

It is equivalent to 1 − *specificity*. The y-axis is TP percentage or recall, which calculated by the formula,

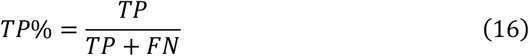

By plotting the TP percentage over FP percentage, ROC curve provides a visual representation of the trade-off between the benefits (TP%) and the costs (FP%).

### 4.2 Experiment one: Evaluate the Overall Performance of AIMDP Model

To demonstrate the effectiveness of AIMDP model, The overall precision, accuracy, and sensitivity are calculated and the results obtained compared to the result obtained from CorrCT[11], COVNet [10], DeConNet[9] and ReNet+[12], as shown in figure 9. To verify the performance of AIMDP through comparisons with other algorithms, the number of images and threshold values are set to the same in all simulations.

**Figure 9:**
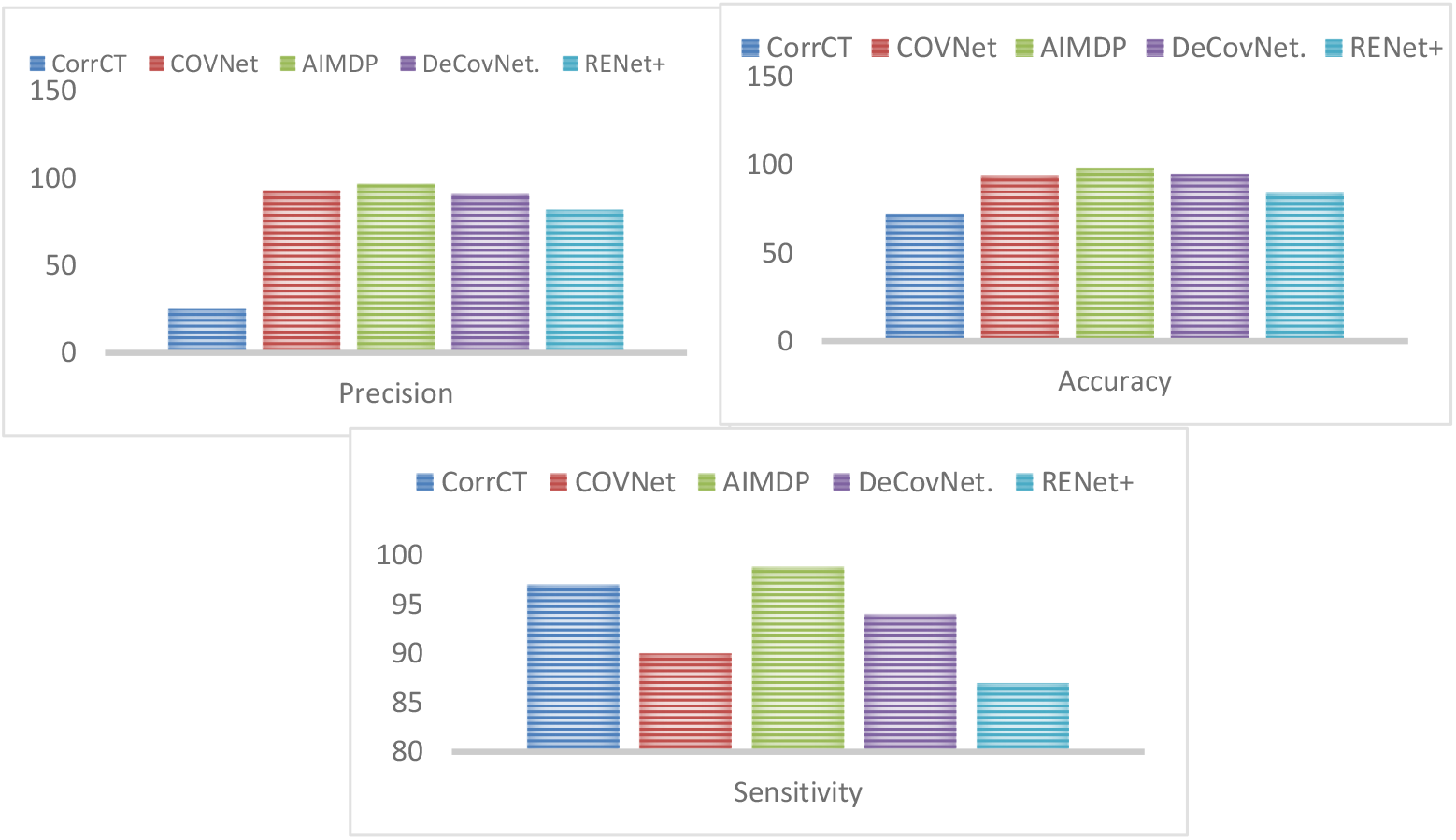
The Overall Precision, Accuracy, and Sensitivity for AIMDP Model Compared to other Models.

Figure 9 reveals the significant superiority of AIMDP over the other models in terms of precision, accuracy, and sensitivity. In addition, figure 10 shows that AIMDP has the lower execution time needed compared to CorrCT[11], COVNet [10], DeCon-Net[9] and ReNet+[12].

**Figure 10:**
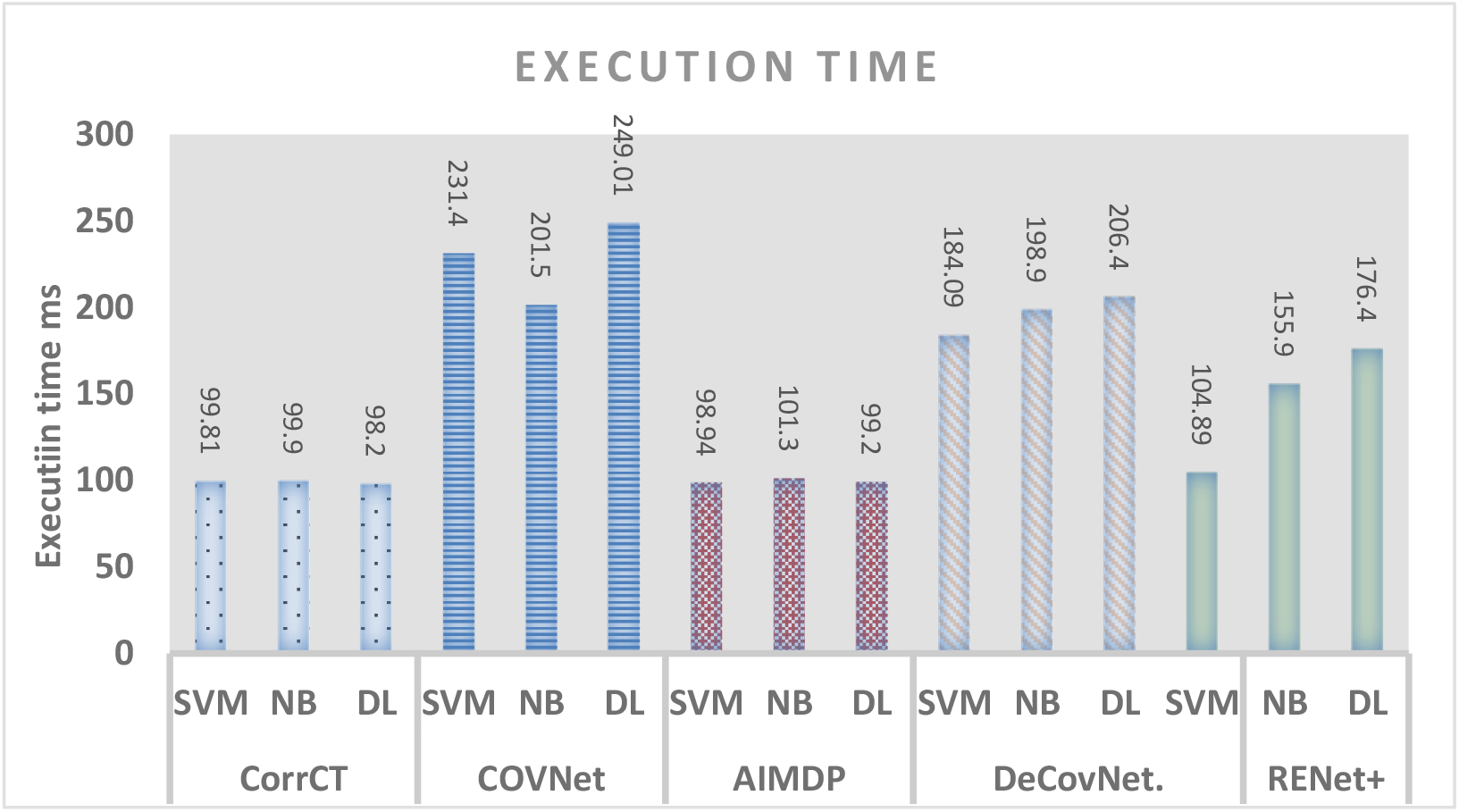
The Execution Time for AIMDP Model Compared to other Models when using different classifiers.

This is due to the presence of many processes included in their executions which may need longer time. Moreover, using the deep learning methods in diagnosing is time consuming. While AIMDP only consider the lung regions and remove the noise in the non-lung regions in the pre-processing phase to avoid time consuming in segmenting the whole image using CNNs.

### 4.3 Experiment 2: Test the PPPV, NPV and ACC values when varying the threshold

This experiment main goal is to test the influence of varying the probability threshold on the prediction and diagnosis process of AIMDP by calculating the PPV: Positive Prediction Value, NPV: Negative Prediction Value and ACC: Accuracy. Figure 11 shows the diagnosing performance for COVID-19 cases by changing the probability thresholds.

**Figure 11.**
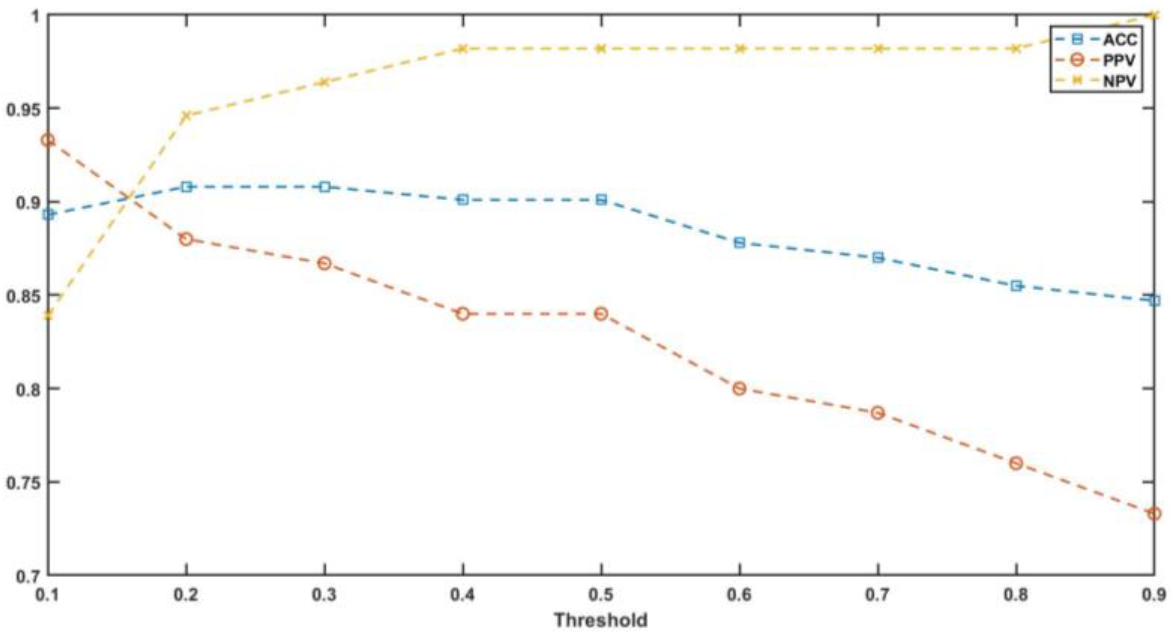
The AIMDP diagnosing performance for COVID-19 cases by changing the probability thresholds.

As shown from figure 11, when the threshold ranged from 0.25 to 0.5, the average value of ACC reached 0.95. while the average value of NPV is 0.97 and PPV is 0.87. that means that the best threshold used must be within this range.

### 4.4 Experiment 3: Test the influence of FSWOA module on AIMDP performance

The feature selection phase is the most critical phase in AIMDP model. WOA algorithm is implemented in this phase to reduce the number of non-relevant features from being processed by AIMDP, which leads to increase the accuracy of the model. FSWOA effect is evaluated in this experiment, as shown in figure 12. The overall precision, recall and accuracy of AIMDP model is tested with and without implementing FSWOA module.

**Figure 12.**
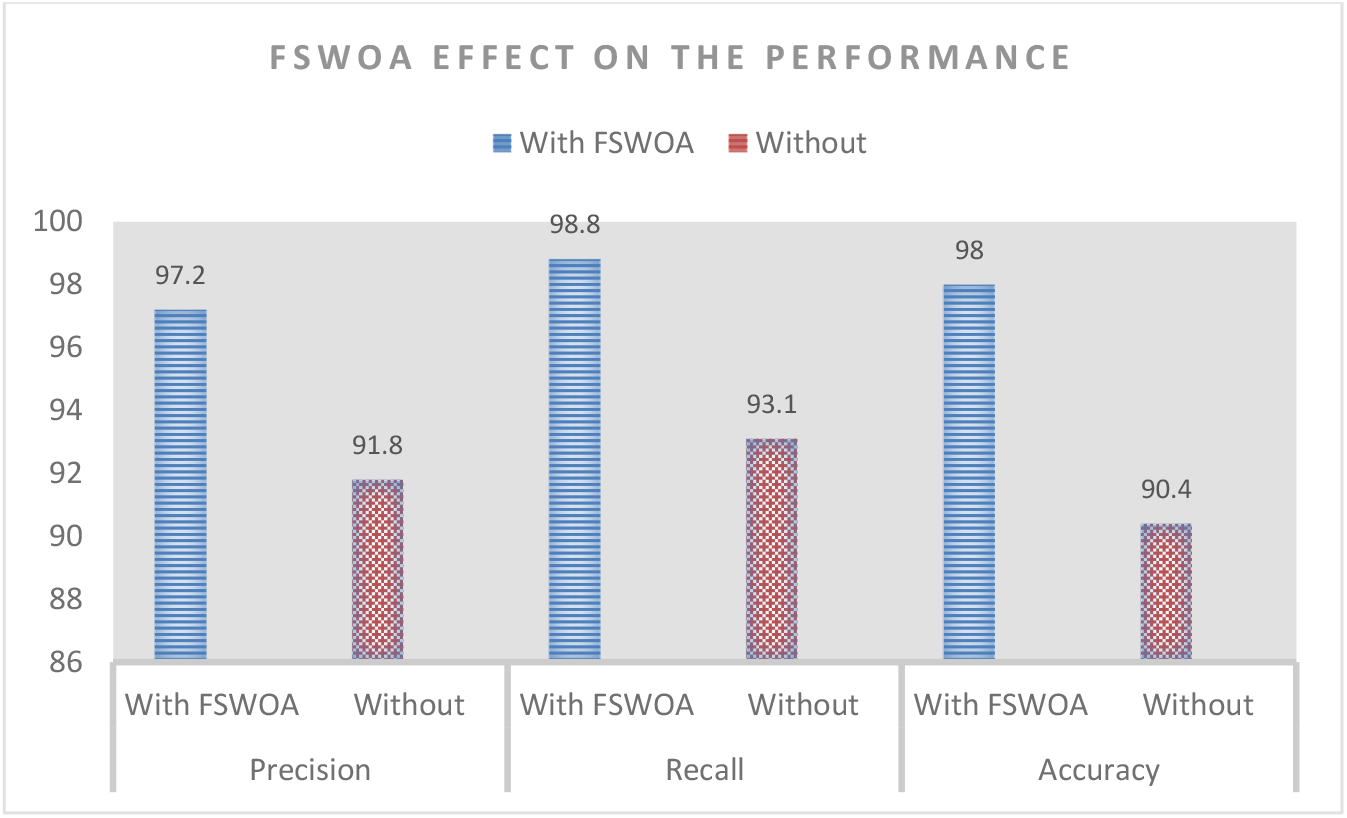
The overall precision, recall and accuracy of AIMDP model with and without implementing FSWOA module.

The performance of AIMDP model when using FSWOA module achieved significant better values compared with the performance of AIMDP without this module

### 4.5 Experiment 4: The Diagnosis Phase Evaluation

The performance of AIMDP during diagnosing COVID-19 patient and other viral pneumonias patient are evaluated using the ROC curve, as shown in figure 13. The sensitivity, specificity and AUC for COVID-19 are 90, 96, 0.96, respectively. The performance for other viral pneumonias yields 94 % for the sensitivity, 96 for the specificity and .98 for AUC.

**Figure 13.**
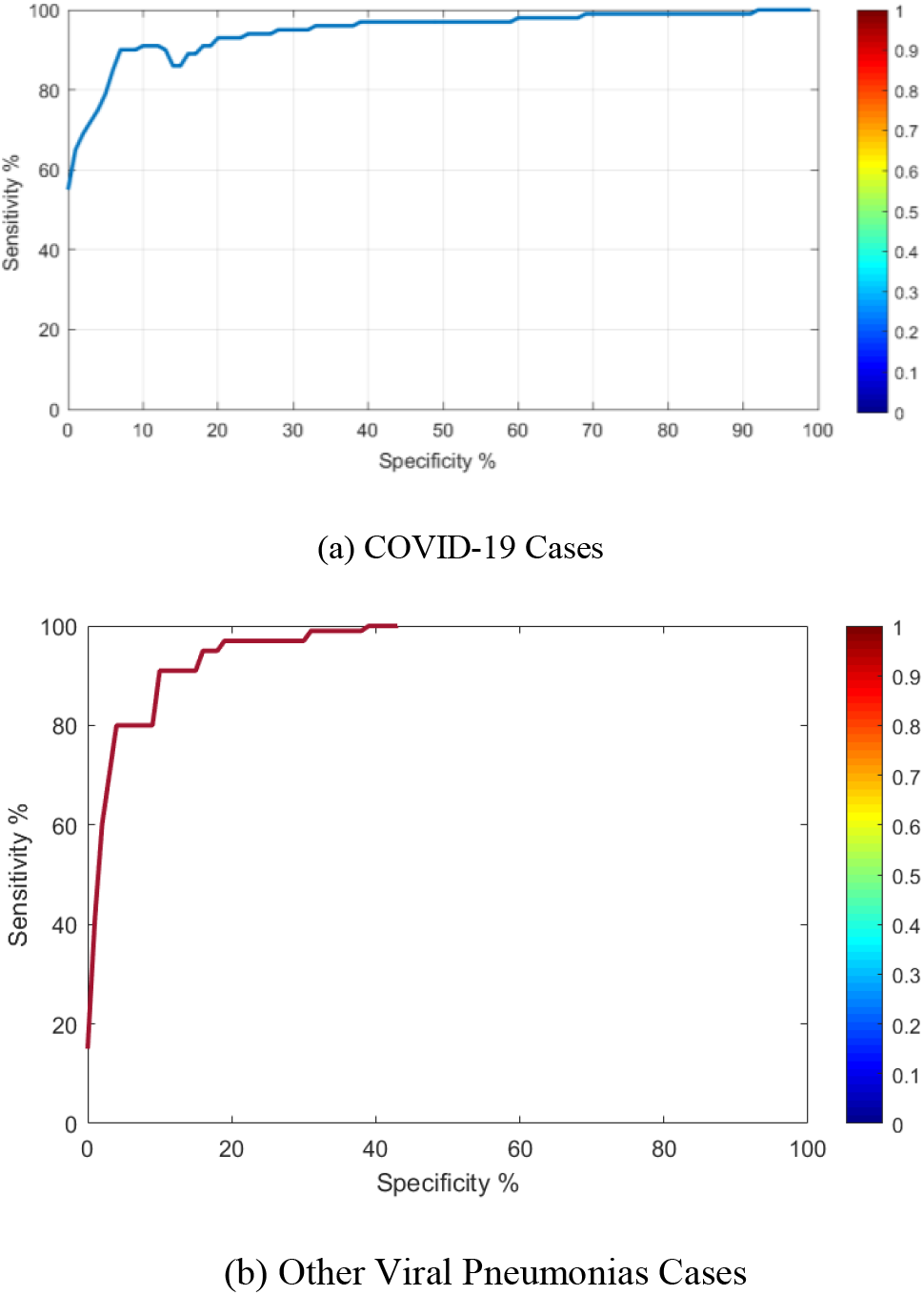
The ROC curves of diagnosing module in AIMDP using testing set for (a) COVID-19 and (b) Other Viral Pneumonias Cases

**Figure 14:**
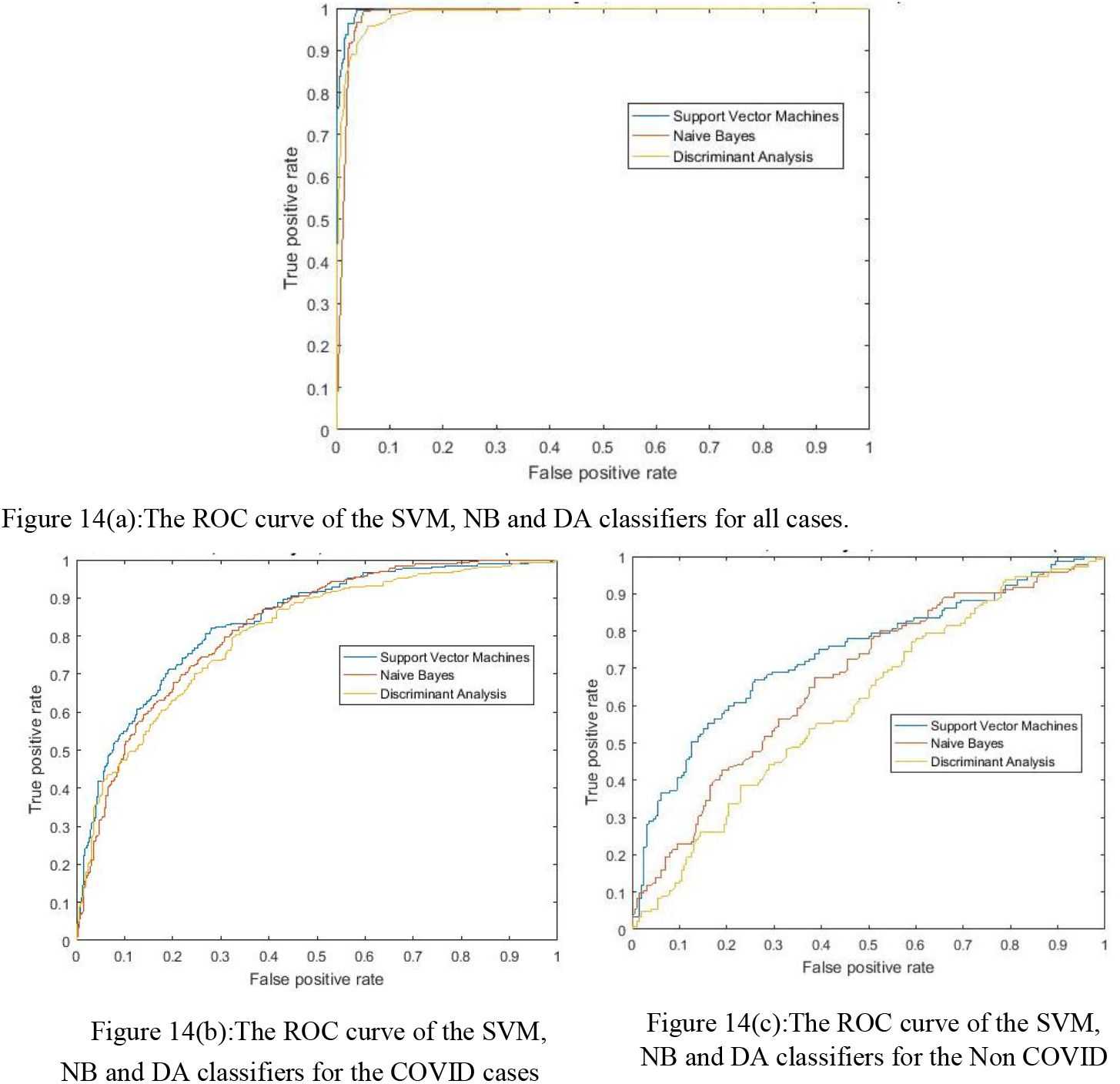
The ROC curve for the three classifiers used in the prediction phase of non-responding cases.

### 4.6 Experiment 5: Test the Prediction Accuracy using different Classifiers

This experiment evaluates the significance and accuracy of prediction phase of AIMDP using different classifiers for prediction. The predictions are compared with random predictions. The standard Predictions (P) value is the percentage of number of random predictions with higher accuracy than the calculated predictions. The performance of three classifiers (SVM, NB and, DA) is tested on the dataset with relevant features extracted from FSWOA phase. The overall accuracy, F-score, G-mean, and the area under the ROC curve (AUC) are evaluated for each classifier, as shown in table 2.

The results indicate that the performance of the SVM classifier is the highest with a value of 97.14 % followed by the NB with a value of 95.99%, then the DA one with a value of 94.1%.

Considering the SVM, the F-score value is 96.84% meaning that both TP and TN are high, simultaneously. Also, the G-mean value (96.93 %) is high, which means that both TPR and TNR are high; thus, indicating that the overall classification performance of the imbalanced dataset is good. This fact is emphasized by the high value of the AUC (0.9957). A Sensitivity score of the COVID of 96.27% indicates the portion of the COVID that is predicted correctly. Likewise, a Specificity score of 97.6% indicates the portion of the Non COVID sample that is predicted correctly, (447 out of 458). This indicates that the SVM is more accurate in predicting the Non COVID in this case. When it comes to the performance of the NB classifier, it is seen that the F-score value is lower than that of the G-mean; however, both are relatively high (above 95.63 %) indicating the goodness of the performance. A high AUC value of 0.9857 confirms an effectiveness of performance. A sensitivity of 97.51% and 95.29 % for the COVID and Non COVID, respectively, indicate that the NB predicts the COVID class more effectively than Non COVID (for this dataset), the contrary of the SVM. As for the DA classifier, the F-score and the G-mean values are close to each other (around 94%), which gives indication that the DA performance is compatible.

However, the sensitivity of the COVID and the Non COVID are 91.29% and 96.51%, respectively, indicating that the DA prediction of the COVID is less effective than that of the Non COVID. The ROC curve of the three classifiers used in the prediction phase of non-responding cases are displayed in the figure 10.

## 5 Conclusions and Future Work

COVID-19 disease has been announced as a pandemic by WHO organization. It leads to thousands of death in short period of time. Accurate and rapid diagnosis of COVID-19 suspected cases plays an essential role in its containment. An Artificial Intelligence-inspired Model for COVID-19 Diagnosis and Prediction for Patient Response to Treatment (AIMDP) is presented. Different AI techniques have been utilized in this model. CNNs is used as a deep learning technique for segmenting the CT images. WOA algorithm is used in the Feature selection phase to select the minimum set of relevant features of diagnosing COVID-19 to optimize the results obtained. An intelligent classifier selector is implemented to select, from three different classifiers, the most appropriate classifier based on the classification error obtained for each case. An automatic diagnosing phase is implemented to accurately distinguish COVID-19 from other viral pneumonias cases, using CT scan images with the most relevant signs of COVID-19 cases and further lab evaluation (RT-PCR and CBC). An adaptive prediction phase is presented to predict the patient ability to respond to the COVID-19 treatment based on different inputs given for the patient.

To the best of our knowledge, this is the first study to perform prediction on Patient response to COVID-19 treatment. This will lead to saving number of resources and efforts by providing the treatment to non-responding patients. And will help decision makers to search for new treatment for non-responders and give more chance to save the responding cases with appropriate treatment. Number of experiments were performed to validate AIMDP performance. The obtained results proved the promising performance of AIMDP in diagnosing and predicting COVID-19 when compared to recent diagnosing and predicting models. AIMDP achieves high AUC and PPV, and low NPV and execution time, compared to others. As a future work, a plan is made to detect the severity degree of COVID-19 cases to provide appropriate treatment and isolation. Moreover, we plan to collect additional CT scans from different centres to evaluate its performance.

## Data Availability

The CT images dataset firstly collected from https://github.com/UCSD-AI4H/COVID-CT, https://www.sirm.org/en/, and https://github.com/ieee8023/covid-chestxray-dataset. Then the CT images are segmented, updated and analyzed to generate a new dataset which is available from the corresponding author on reasonable request.

https://github.com/UCSD-AI4H/COVID-CT

https://www.sirm.org/en/

